# Observational assessments of the relationship of dietary and pharmacological treatment on continuous measures of dysglycemia over 24 hours in women with gestational diabetes

**DOI:** 10.1101/2022.09.16.22280027

**Authors:** Cassy F. Dingena, Melvin J. Holmes, Matthew D. Campbell, Janet E. Cade, Eleanor M. Scott, Michael A. Zulyniak

## Abstract

**Objectives:** Studies that use continuous glucose monitoring (CGM) to monitor women with gestational diabetes (GDM) highlight the importance of managing dysglycemia over a 24-hour period. However, the effect of current treatment methods on dysglycemia over 24-hrs are currently unknown. This study aimed to characterise CGM metrics over 24-hrs in women with GDM and the moderating effect of treatment strategy.

**Methods:** Retrospective analysis of CGM data from 128 women with GDM in antenatal diabetes clinics. CGM was measured for 7-days between 30-32 weeks gestation. Non-parametric tests were used to evaluate differences of CGM between periods of day (morning, afternoon, evening, and overnight) and between treatment methods (i.e., diet alone or diet+metformin). Exploratory analysis in a subgroup of 34 of participants was performed to investigate the association between self-reported macronutrient intake and glycaemic control.

**Results:** Glucose levels significantly differed during the day (i.e., morning to evening; P<0.001) and were significantly higher (i.e., mean blood glucose and AUC) and more variable (i.e., SD and CV) than overnight glucose levels. Morning showed the highest amount of variability (CV; 8.4% vs 6.5%, P<0.001 and SD; 0.49 mmol/L vs 0.38 mmol/L, P<0.001). When comparing treatment methods, mean glucose (6.09 vs 5.65 mmol/L; P<0.001) and AUC (8760.8 vs 8115.1 mmol/L.hr; P<0.001) were significantly higher in diet+metformin compared to diet alone. Finally, the exploratory analysis revealed a favourable association between higher protein intake (+1SD or +92 kcal/day) and lower mean glucose (-0.91 mmol/L p, P=0.02) and total AUC (1209.6 mmol/L.h, P=0.021).

**Conclusions:** Glycemia varies considerably across a day, with morning glycemia demonstrating greatest variability. Additionally, our work confirms that individuals assigned to diet+metformin have greater difficulty managing glycemia and results suggest that increased dietary protein may assist with management of dysglycemia. Future work is needed to investigate the benefit of increased protein intake on management of dysglycemia.

## Introduction

Pregnancy induces a natural state of insulin resistance (IR) to shuttle a greater proportion of maternal nutrients to the infant for growth and development (1). However, in 5-18% of all UK pregnancies (2, 3) this metabolic shift leads to uncontrolled and unhealthy increases in blood glucose (1, 4-6), known as gestational diabetes mellitus (GDM). GDM occurs when women not previously known to have diabetes develop hyperglycemia during pregnancy, risking the health of mother and growing offspring (5, 7). Moreover, GDM is associated with increased risk of pre-eclampsia, preterm delivery, and type 2 diabetes (T2DM) in later life (8); while offspring exposed to GDM in utero are at increased risk of abnormal birth weight, birth injury, mortality, and obesity and T2DM in later life (7-9). Treatment aims to control maternal glucose levels and mitigate adverse pregnancy outcomes and long-term maternal and offspring health risks (10).

The first line of treatment for GDM typically consists of dietary and lifestyle education (1, 11). Diets focussing on low glycaemic index (GI) foods and reduced overall carbohydrate intake are most common for the management of GDM(1, 3) but no consensus on the best nutritional approach has been agreed (12, 13). In the UK, clinical recommendations focus on improving carbohydrate quality and reducing overall carbohydrate intake (3, 6). While replacing simple carbohydrates with higher-quality carbohydrates and lower overall carbohydrate intake can help to control glucose levels, its effectiveness on managing dysglycemia is not consistent between populations (13), with meta-analyses demonstrating high levels of heterogeneity (>60%) of low GI diets on fasting and post-prandial glucose levels (14). This may be because trials often prescribe specific low-GI nutrients to be consumed at defined times over a 24-hour period, while real-life meals are often mixtures of foods consumed at various points throughout the day (15-17). Previous research has demonstrated that dietary protein can attenuate the subsequent rise in the postprandial glucose response (PPGR) (18, 19). However, free living individuals consume meals that consist of mixed macronutrients consumed at different times of the day, suggesting that a single measure of post-prandial glucose (PPG) may be inadequate to characterise the full effect of diet on dysglycemia.

Randomised controlled trials suggest that 80% of women with GDM can achieve normal glucose levels through diet and lifestyle modification alone (20). However, where management of dysglycemia is more difficult, pharmacological therapy may be needed. Metformin, an oral antihyperglycemic drug, has been used as a secondary line treatment for glycemic control in T2DM for decades (21, 22). In women with GDM, the UK clinical guidelines also recommend metformin as secondary-line treatment in the management of dysglycemia (3), with added benefits linked to reduced gestational weight gain, maternal hypertensive disorders, macrosomia, neonatal hypoglycemia, and intensive care unit admissions (3). Current evidence suggests no difference in standard maternal measures of glycaemia or neonatal outcomes after delivery in women treated with either diet or metformin (23).

However, maternal glucose is dynamic, glucose tolerance and insulin sensitivity vary over a 24-hour period (24, 25), and emerging evidence suggests that glycaemic spikes and patterns rather than single measures of glycaemia may be more indicative of poor dysglycemic management and provide novel information regarding maternal and offspring health risks (26). These details are captured using continuous glucose monitors (CGM), which repeatedly record glucose measures in close succession (minutes) over a specific period of time (days or weeks), and offer detailed records of glucose dynamics (27). The capabilities of CGM recently demonstrated novel associations between CGM-defined markers of dysglycemia at (i) 12-weeks’ gestation with infant health outcomes [i.e., preterm birth: OR = 1.52 (1.08, 2.13); large-for-gestational age: OR = 1.49 (1.06, 2.08)] and (ii) 24 -week gestation with maternal outcomes [pre-eclampsia: OR = 1.98 (1.17, 3.37)] (28). This suggests that CGM can (i) offer new information regarding the association between dysglycemia, and maternal and offspring health, and (ii) be used to inform and direct care more accurately and at an earlier point of pregnancy. Interestingly, CGM has not yet been used to evaluate the relationship between lifestyle treatment with or without metformin to glucose spikes and variability over a 24-hour period in women with GDM, which could offer novel insights regarding treatment strategies (i.e., diet or diet+metformin) as mediators of dysglycemia across the day in GDM pregnancies. Therefore, this study aimed to determine key time points during the day of disrupted glucose control, and the relationship of treatment and dietary mediators to this disrupted glucose control in a diverse population of pregnant women with GDM.

## Methods

### Study Design

Secondary retrospective analysis of an observational cohort of 162 pregnant women with GDM (2). Of 162 women, 128 had complete participant data and < 30% missing CGM data across the 7 days (**Supplementary Figure 1**). CGM data was collected between 16/01/2014 and 23/08/2016. All women provided written informed consent. The study was approved by the Yorkshire and Humber Regional Ethics Committee (13/YH/0268) and NHS Health Research Authority (NRES) Committee South Central–Oxford C (14/SC/1267).

### Study Participants

Participants were between 18 and 45 years of age, had a singleton pregnancy, recruited from antenatal diabetes clinics in Leeds Teaching Hospitals Trust and were diagnosed with GDM according to National Institute for Health and Care Excellence (NICE) guideline criteria — i.e., fasting glucose ≥5.6 mmol/L (≤100.8 mg/dL) and/or 2-h glucose ≥7.8 mmol/L (≥140.4 mg/dL) after a 75-g oral glucose tolerance test at ∼26 weeks of gestation (3). As per clinical guidelines, all women were advised to aim for self-monitored blood glucose (SMBG) targets: fasting glucose <5.3 mmol/L and 1-h post meal <7.8 mmol/L (2, 28). Women were treated with diet and lifestyle modifications as first-line therapy and with metformin and/or insulin as second-line therapy. NICE guidelines state that if blood glucose targets are not achieved with diet and lifestyle changes within 1 to 2 weeks, metformin will be offered(3). All women with GDM attending the antenatal diabetes clinic at Leeds Teaching Hospital Trust were invited to participate. Exclusion criteria included having a physical or psychological disease likely to interfere with the conduct of the study, and not speaking English.

### Continuous glucose monitoring (CGM)

The CGM device used was iPro2 (Medtronic). The CGM data was calibrated by simultaneous SMBG using approved and standardized blood glucose meters and test strips (Contour XT; Bayer) (26). Data was anonymised using a unique identification number for each participant and was downloaded via CareLink (Medtronic) for analysis. The device measures glucose levels every 5 minutes over a 24-hour period, providing 288 measures every day for 7 days. To analyse mean glycemic control over a 24-hr period, the individual timepoint measurements were averaged across 7 days. This provided 288 average measures of glucose over a 24-hr period.

To analyse key time points across the 24-hr day, the CGM glucose data was analysed by dividing the data into four equal periods of six hours (e.g., morning 06:00-11:55, afternoon 12:00-17:55, evening 18:00-23.55, and overnight 00:00-05.55). These windows were chosen so that the morning, afternoon, and evening time periods include pre- and post-prandial glucose levels, and the overnight time-period monitors a sleep cycle and a sustained fasted state. To evaluate dysglycemia, our primary outcome of interest was coefficient of variation (CV). However, additional indices were examined for the full 24hr hours and for each period, including: mean glucose levels, standard deviation (SD), and the percentage of time spent within the pregnancy glucose target range (TIR; 3.5–7.8 mmol/L [70.2–140.4 mg/dL]), time spent above (TAR; >7.8 mmol/L [≥140.4 mg/dL]) and below (TBR; <3.5 mmol/L [≤70.2 mg/dL]) target range(27).

### Nutritional data

In an exploratory analysis, complete nutritional information was available in a subgroup of 34 of the 128 women with CGM data (Supplementary figure 1). Average daily dietary intake was collected using an online food diary (myfood24)(29). Participants were instructed to complete the online record for 5 days. Dietary intake was recorded as mean total grams or kilocalories per day. After removal of 1 participant with an implausible total kilocalorie intake <500 kcal/day (30), the nutrient residual model was used to perform tests for linear association between individual macronutrients and glycemic measures in 33 participants (31), after adjustment for maternal age, ethnicity, parity, maternal BMI, and weeks of gestation (32, 33). Briefly, the nutrient residual model reduces confounding by using the residuals of total energy intake, which represent the difference between each individual’s actual intake and the intake predicted by their total energy intake, thereby removing the variation caused by total energy intake rather than absolute intake (31). Total kilocalorie intake per day for each participant was standardised to the average energy intake per day within our study (1500 kcal/day). To assess the association of macronutrients and glycemic control, we constructed multiple variable regression models for each CGM metric (e.g., mean glucose, SD, CV, AUC, iAUC, TIR, TAR or TBR). Each model CGM model included all macronutrients— i.e., total carbohydrate intake (kcal) + total fat intake (kcal) + total energy intake (kcal) — and covariates (maternal age, ethnicity, parity, maternal BMI, and weeks of gestation). This model permits the assessment of substituting carbohydrates, fats, or proteins (reflected by total energy intake) with an isocaloric equivalent quantity of the other macronutrients. Specifically, these models examine the association of each macronutrient independently with CGM metrics, when all other variables (i.e., other macronutrients, energy, and covariates) are held constant. With three macronutrient sources of energy, when ‘carbohydrates’ and ‘fats’ are held constant, the increase in the ‘calorie’ variable represents an increase in ‘protein’ (31).

### Statistical analysis

Friedman’s test and pairwise Wilcoxon signed rank test were used because of visually apparent asymmetric data, with Bonferroni corrections applied for multiple comparisons between periods of the day. Recent evidence suggests a difference in effect size of 0.924 (Cohen’s d) on mean glucose between diet and diet+metformin; therefore, at 80% power we required ≥ 21 participants between comparison groups (34). To assess the association between dietary macronutrients and glycaemic control, multiple variable linear regression analyses were performed and adjusted for maternal age, ethnicity, parity, maternal BMI, and gestational week. The Cook’s Distance was used for influential outlier assessment. Statistical significance was set at p<0.05. All statistical analyses were conducted in RStudio (version 4.0.3), and all figures were created in GraphPad Prism 9.

## Results

Over a 24-hour period, glucose measures were collected every 5 minutes, yielding a total of 288 glucose measurements per individual and a total of 36,864 glucose measurements for 128 women. In total, 34 women were excluded, due to incomplete participant data and <30% missing CGM data across the 7 days. The majority of participants self-identified as white European (61%) and managed their dysglycemia with diet alone (n=58), diet+metformin (n=51), diet+insulin (n=2), or diet+metformin+insulin (n=17). Due to small numbers and inadequate power of insulin and metformin+insulin treatment groups (i.e., <21 participants), analysis on treatment effect was limited to diet and diet+metformin groups. The average age and BMI of participants was 33 years and 30.6 kg/m^2^. Approximately 30% of women, 34 out of 128 with available CGM data, used myfood24 to record their dietary intake. Participant characteristics are summarised in **Table 1**.

**Table 1.**
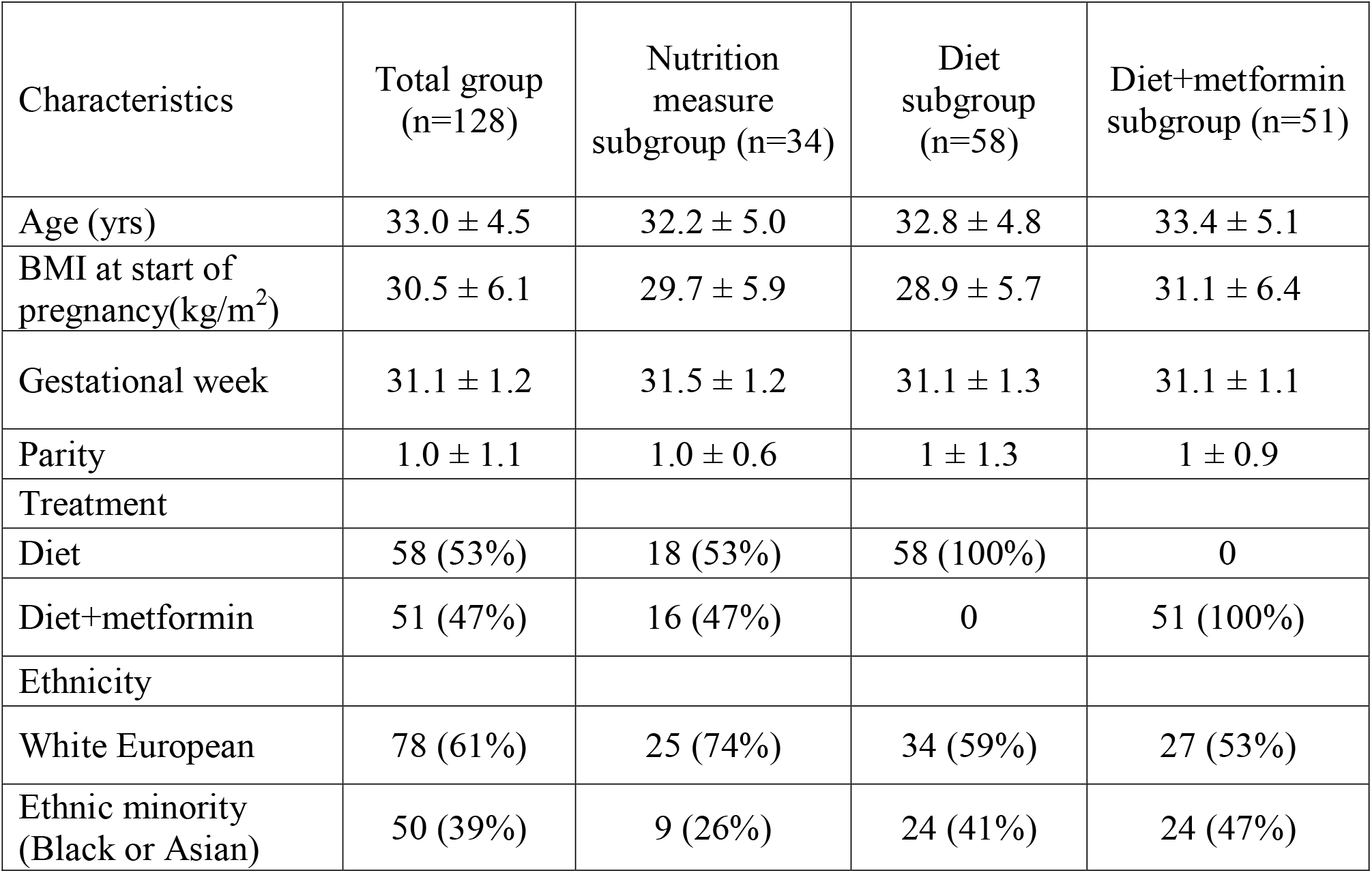
Participant characteristics

| Characteristics | Total group<br>(n=128) | Nutrition<br>measure<br>subgroup (n=34) | Diet<br>subgroup<br>(n=58) | Diet+metformin<br>subgroup (n=51) |
| --- | --- | --- | --- | --- |
| Age (yrs) | 33.0 ± 4.5 | 32.2 ± 5.0 | 32.8 ± 4.8 | 33.4 ± 5.1 |
| BMI at start of<br>pregnancy(kg/m<sup>2</sup>) | 30.5 ± 6.1 | 29.7 ± 5.9 | 28.9 ± 5.7 | 31.1 ± 6.4 |
| Gestational week | 31.1 ± 1.2 | 31.5 ± 1.2 | 31.1 ± 1.3 | 31.1 ± 1.1 |
| Parity | 1.0 ± 1.1 | 1.0 ± 0.6 | 1 ± 1.3 | 1 ± 0.9 |
| Treatment |  |  |  |  |
| Diet | 58 (53%) | 18 (53%) | 58 (100%) | 0 |
| Diet+metformin | 51 (47%) | 16 (47%) | 0 | 51 (100%) |
| Ethnicity |  |  |  |  |
| White European | 78 (61%) | 25 (74%) | 34 (59%) | 27 (53%) |
| Ethnic minority<br>(Black or Asian) | 50 (39%) | 9 (26%) | 24 (41%) | 24 (47%) |

### CGM analysis

An effect of “time of day” was identified for the majority of CGM metrics — including, mean glucose, SD, CV, AUC, iAUC, and TAR (**Figure 1 and Table 2**). Therefore, pairwise analyses were performed on all CGM metrics. For CV and SD, measures were relatively stable during the day but lowered ‘overnight’ (Figure 1). Conversely, glucose and total AUC increased steadily from morning to evening and dropped overnight (mean glucose and AUC; all time comparisons P>0.001). When focussing on measures of glycemic variability, SD and CV of glucose were greatest in the morning and steadily decreased towards the lowest levels overnight (SD; 0.49mmol/L vs 0.30mmol/L and CV; 8.41% vs 4.99%, P<0.001). iAUC fluctuated over the 24-hour period, with the highest levels recorded in the morning and evening (1244.5 vs 1311.6 mmol/L.min^-1^, P=0.87), reductions in the afternoon (1106.0 mmol/L.min^-1^, P<0.001) and recording the lowest levels overnight (604.9 mmol/L.min^-1^, P<0.001). The Friedman test reported no significant differences when glucose levels were within (TIR), or below (TBR) a specific range, no differences were confirmed between times-of-day either (Figure 1 and Table 2). However, TAR significantly differs across the day and was highest during the evening (TAR evening; 4.41%, P=0.018).

**Figure 1.**
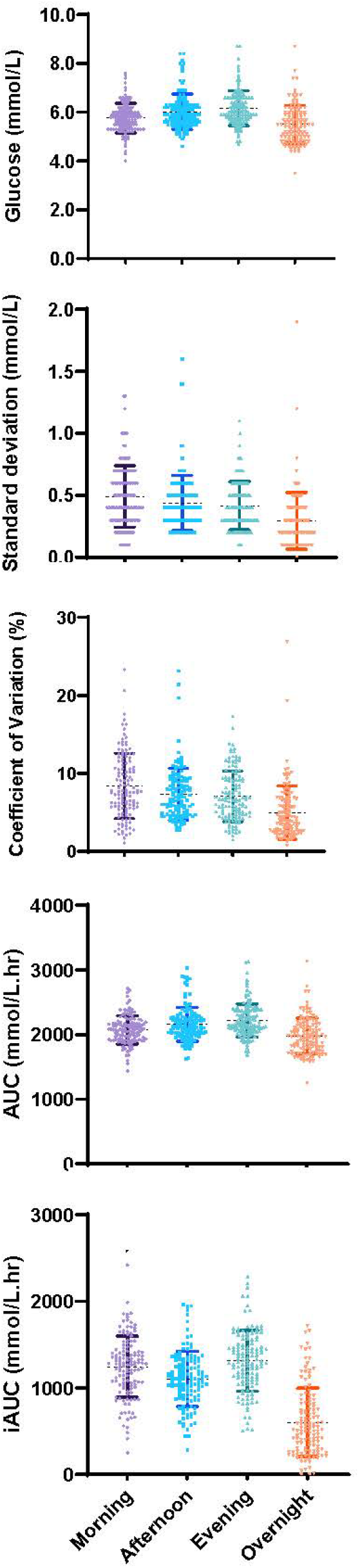
Mean measures of continuous glucose monitoring CGM across distinct periods of a for 128 women with GDM.

**Table 2.**
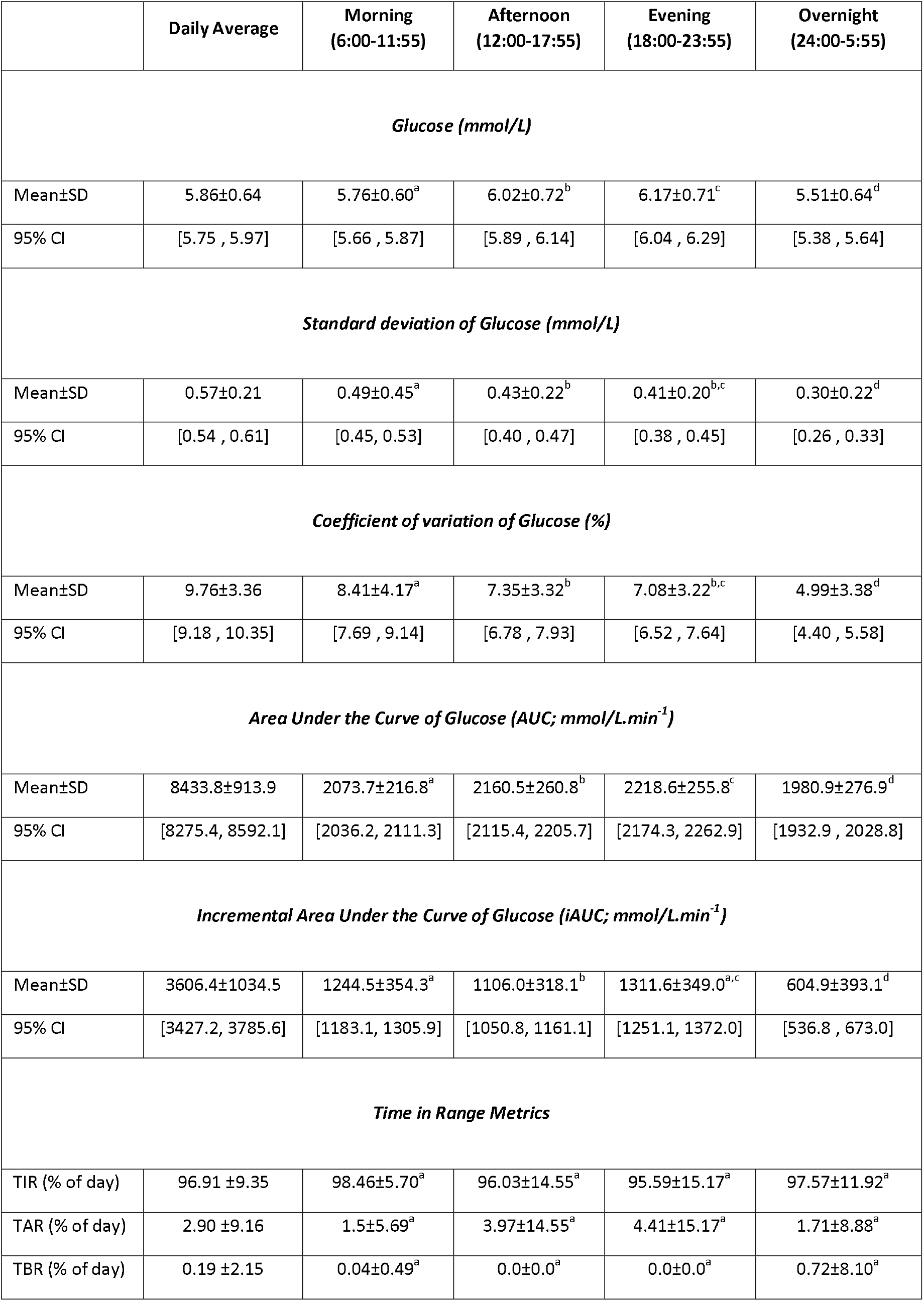
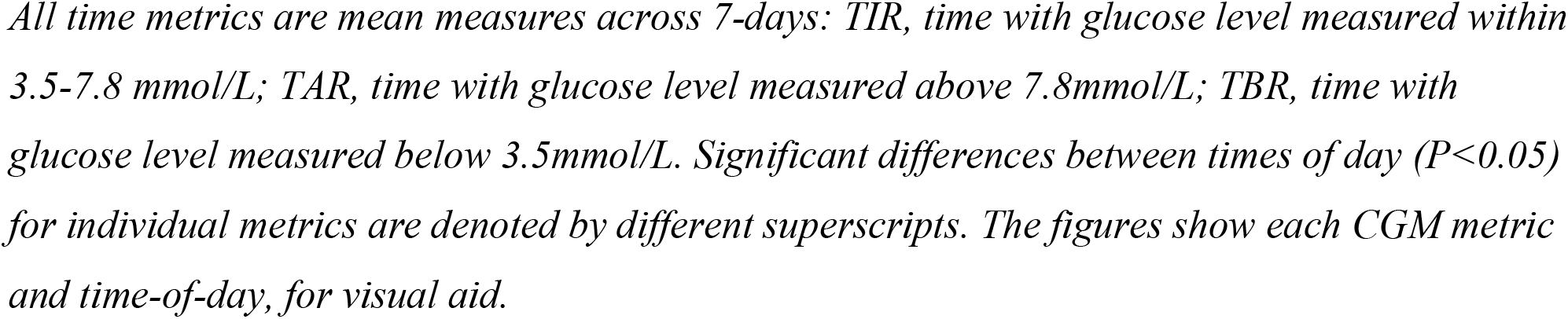
Summary of measures of continuous glucose monitoring CGM over a 24-hour period.

|  | Daily Average | Morning<br>(6:00-11:55) | Afternoon<br>(12:00-17:55) | Evening<br>(18:00-23:55) | Overnight<br>(24:00-5:55) |
| --- | --- | --- | --- | --- | --- |
| *Glucose (mmol/L)* |  |  |  |  |  |
| Mean±SD | 5.86±0.64 | 5.76±0.60<sup>a</sup> | 6.02±0.72<sup>b</sup> | 6.17±0.71<sup>c</sup> | 5.51±0.64<sup>d</sup> |
| 95% CI | [5.75 , 5.97] | [5.66 , 5.87] | [5.89 , 6.14] | [6.04 , 6.29] | [5.38 , 5.64] |
| *Standard deviation of Glucose (mmol/L)* |  |  |  |  |  |
| Mean±SD | 0.57±0.21 | 0.49±0.45<sup>a</sup> | 0.43±0.22<sup>b</sup> | 0.41±0.20<sup>b,c</sup> | 0.30±0.22<sup>d</sup> |
| 95% CI | [0.54 , 0.61] | [0.45, 0.53] | [0.40 , 0.47] | [0.38 , 0.45] | [0.26 , 0.33] |
| *Coefficient of variation of Glucose (%)* |  |  |  |  |  |
| Mean±SD | 9.76±3.36 | 8.41±4.17<sup>a</sup> | 7.35±3.32<sup>b</sup> | 7.08±3.22<sup>b,c</sup> | 4.99±3.38<sup>d</sup> |
| 95% CI | [9.18 , 10.35] | [7.69 , 9.14] | [6.78 , 7.93] | [6.52 , 7.64] | [4.40 , 5.58] |
| *Area Under the Curve of Glucose (AUC; mmol/L.min<sup>-1</sup>)* |  |  |  |  |  |
| Mean±SD | 8433.8±913.9 | 2073.7±216.8<sup>a</sup> | 2160.5±260.8<sup>b</sup> | 2218.6±255.8<sup>c</sup> | 1980.9±276.9<sup>d</sup> |
| 95% CI | [8275.4, 8592.1] | [2036.2, 2111.3] | [2115.4, 2205.7] | [2174.3, 2262.9] | [1932.9 , 2028.8] |
| *Incremental Area Under the Curve of Glucose (iAUC; mmol/L.min<sup>-1</sup>)* |  |  |  |  |  |
| Mean±SD | 3606.4±1034.5 | 1244.5±354.3<sup>a</sup> | 1106.0±318.1<sup>b</sup> | 1311.6±349.0<sup>a,c</sup> | 604.9±393.1<sup>d</sup> |
| 95% CI | [3427.2, 3785.6] | [1183.1, 1305.9] | [1050.8, 1161.1] | [1251.1, 1372.0] | [536.8 , 673.0] |
| *Time in Range Metrics* |  |  |  |  |  |
| TIR (% of day) | 96.91 ±9.35 | 98.46±5.70<sup>a</sup> | 96.03±14.55<sup>a</sup> | 95.59±15.17<sup>a</sup> | 97.57±11.92<sup>a</sup> |
| TAR (% of day) | 2.90 ±9.16 | 1.5±5.69<sup>a</sup> | 3.97±14.55<sup>a</sup> | 4.41±15.17<sup>a</sup> | 1.71±8.88<sup>a</sup> |
| TBR (% of day) | 0.19 ±2.15 | 0.04±0.49<sup>a</sup> | 0.0±0.0<sup>a</sup> | 0.0±0.0<sup>a</sup> | 0.72±8.10<sup>a</sup> |

*All time metrics are mean measures across 7-days: TIR, time with glucose level measured within 3.5-7.8 mmol/L; TAR, time with glucose level measured above 7.8mmol/L; TBR, time with glucose level measured below 3.5mmol/L. Significant differences between times of day ( $P<0.05$ ) for individual metrics are denoted by different superscripts. The figures show each CGM metric and time-of-day, for visual aid.*

### Exploratory Analysis

#### Treatment Data

Our exploratory post-hoc analysis of treatment included 109 women (n=58 in diet subgroup and n=51 in diet+metformin). A significant association of treatment on mean glucose and AUC was found (F (3,1)=27.3, P<0.001 and F(3,1)=28.9, p<0.001, respectively), both mean glucose (5.65 vs 5.97mmol/L) and total AUC (8115.1 vs 8586.1 mmol/L.min^-1^) was higher in metformin subgroup. No interaction between time-of-day and treatment on CGM metric was found.

#### Nutrient Data

Our exploratory analysis of nutritional data included 34 women (**Table 3**). Of the 8 CGM metrics assessed, mean glucose and AUC showed significant associations with dietary mediators. To clarify, these models examine the association of each macronutrient with glycemic metrics, when the other macronutrients are held at a constant level — e.g., carbohydrates when intake of dietary fat and protein are held constant. With only three macronutrient sources of energy (i.e., carbohydrates, fats, and protein), when ‘carbohydrates’ and ‘fats’ are held constant, any increase in the ‘calorie’ variable represents an increase in ‘protein’ (31). After adjusting for known confounders (i.e., maternal age, BMI, gestational age at CGM measurement, parity, ethnicity, and treatment), an increase (+1 SD) of fats or carbohydrates associated with higher mean 24-hr glucose and AUC glucose (**Table 4**), while dietary protein (+1SD) associated with reduced mean 24-hr glucose (-0.91mmol/L; P=0.02) and AUC glucose (-1296 mmol/L.min^-1^; P=0.021).

**Table 3.** Nutritional intake: Average values of nutrients intake reported by random subsample of 39 participants that maintained dietary records.

|  | **Daily intake (kcal/day)**<br>(% total kcal/day) | **Daily intake**<br>(gram/day) |
| --- | --- | --- |
| **Protein** | 246±92<br>(16%) | 61±26 |
| **Fats** | 577±290<br>(38%) | 64±33 |
| **Carbohydrates** | 716±311<br>(47%) | 176±74 |
| *Non-sugar* | 474±208 | 117±50 |
| *Sugar* | 242±179 | 59±43 |
| **Total intake** | 1513±517 | N/A |

*Data reported as mean intake ± standard deviation (SD) per day of each nutrient and total energy intake. Mean proportion of nutrients of total caloric intake reported in parentheses.*

**Table 4.** Multivariable regression of dietary mediators (carbohydrates, fats, and protein) and glycemia stratified by outcome metric of 34 participants that maintained dietary records and had CGM metrics available.

| Variables | Mean glucose (mmol/L) |  | AUC (mmol/L.min<sup>-1</sup>) |  |
| --- | --- | --- | --- | --- |
| | $\beta$ (95% CI) | P-value | $\beta$ (95% CI) | P-value |
| Age | -0.015 (-0.05, 0.02) | 0.38 | -22.1 (-70.2, 25.9) | 0.38 |
| Maternal BMI | 0.022 (-0.005, 0.05) | 0.12 | 31.8 (-7.1, 70.7) | 0.12 |
| Gestational week | 0.009 (-0.12, 0.14) | 0.89 | 12.5 (-173.3, 198.3) | 0.90 |
| Parity | 0.093 (-0.24, 0.28) | 0.49 | 132.5 (-240.4, 505.3) | 0.50 |
| Ethnicity | 0.22 (-0.36, 0.4) | 0.93 | 23.2 (-526.2, 572.6) | 0.93 |
| Treatment type | 0.17 (-0.08, 0.52) | 0.17 | 315.5 (-121.5, 752.5) | 0.17 |
| Adjusted carbohydrates | 0.63 (0.13, 1.1) | **0.021** | 887.9 (173.6, 1602.2) | **0.023** |
| Adjusted fats | 0.49 (0.04, 0.93) | **0.043** | 694.7 (48.5, 1340.8) | **0.046** |
| Adjusted protein | -0.91 (-0.2, -1.6) | **0.02** | -1296.0 (-265.0, -2327.0) | **0.021** |

Mean glucose $r^2 = 0.32$ , AUC $r^2 = 0.18$ . Treatment was coded as follows: 0=diet, 1=diet+metformin. Parity was reported as having 0, 1, 2, 3, 4, 5 or 6 children. Ethnicity was coded as: 0=White and 1=Ethnic minority (e.g., Asian, Black African). CI = confidence interval. Significant associations ( $P < 0.05$ ) in bold.

## Discussion

In an observational cohort of 162 women with GDM, this study demonstrated that (i) CGM offers different methods of assessing glycemic health; (ii) measures of dysglycemia vary considerably over a 24-hour period; and (iii) distinct periods of day are prone to lower or higher levels of absolute glucose as well as glucose variability. Depending on the CGM metric used, ‘morning’ and ‘overnight’ showed to be times of greatest dysglycemia. More specifically, glucose levels were most variable during the day (morning to evening) but were stable in a healthy range (∼95% of the time), while ‘overnight’ showed extended periods of lower glucose levels with relatively less glucose variability. Additionally, exploratory analysis of the association between treatment type (diet vs diet+metformin), time-of-day and maternal glycemic control showed no significant interaction between treatment type and time-of-day on maternal glycemia over a mean 24h period. However, individuals assigned to diet with metformin appeared to have higher levels of dysglycemia, as reflected by elevated mean glucose and total AUC.

Current measures of dysglycemia often use fasting or mean glucose levels to evaluate glycemic control. In our analysis, we report the mean morning, afternoon, and evening glucose levels to be significantly higher compared to mean glucose levels overnight. This agrees with existing understanding of overnight glycemic control, with glucose levels typically falling overnight(35). However, recent work has speculated that glucose excursions quantify a health risk that is independent of mean glucose levels (36, 37). The proposed standard metric for glycemic variability is the CV of glucose (27, 37), which quantifies the magnitude of glycemic variability standardised to mean glucose levels. Despite seeing no difference in mean glucose levels between, afternoon, and evening, our study shows that CV steadily declines during the day reaching lowest values ‘overnight’ and reports that morning CV was significantly higher compared to other times-of-day. This agrees with trends observed in non-diabetic men and women (n=60) that reported significantly higher Daytime CV (06:00-21:59) compared to Overnight CV (22:00-05:59) (38) but disagrees with evidence from adolescent boys and girls (n=107; 13.1 ±2.6 years) that suggests CV increases from early morning (06:00) and peaks from midday to late-night (12:00-23:00) (39). However, the significance in temporal CV patterns was not formally assessed for adolescents, so its importance is uncertain. Recent work suggests that diabetes CV is involved with offspring growth in the 2^nd^ trimester in women with type-1 diabetes (40, 41), and may be an indicator of risk of future health complications associated with T2DM (including cardiovascular disease, coronary events, non-cardiovascular mortality, and total mortality) (4). Therefore, morning control of glucose variability (measured by SD and CV) may be a key point of interest for managing maternal and offspring health. Increased morning CV in this study’s group of women might also be the result of a lack in regular routine, these women may need to get their other children ready for school and/or get ready for work and may not have time for breakfast.

Our exploratory post-hoc analysis of treatment effect demonstrated a significant relationship between treatment group and 2 of the 8 CGM metrics showing persistent higher mean glucose levels and total AUC in women treated with diet+metformin. Despite the lack of a significant relationship between metformin treatment group and other CGM metrics, it is important to note that blood glucose levels vary significantly day by day and glycemic control and variability depend on a variety of different exogenous and endogenous determinants such as, elevated insulin resistance, elevated hepatic glucose production, increased production of antagonistic hormones to insulin, sedentary lifestyle, unhealthy dietary habits and age related metabolic deterioration (42). Although metformin is the most commonly prescribed antihyperglycemic medication for diabetes in the U.K., its effectiveness in glycemic control is only now being documented. Noteworthy, metformin is only prescribed when women are failing to achieve glucose targets with diet alone; therefore, glucose levels in this group are higher. Estimates from recent trials suggest that at higher doses metformin can reduce HbA1c by 1–2% (11–22 mmol/mol)(43), this is promising as it has been reported that a 1 % reduction in HbA1c in women with GDM is associated with improved maternal and offspring outcomes (44). Furthermore, a recent study by Bashir et al (20) found that women with GDM on pharmaceutical treatment were diagnosed earlier than women on dietary treatment, and it is likely that early treatment intensification with diet and metformin has led to reduced foetal glucose levels, foetal hyperinsulinemia and macrosomia.

In our exploratory analysis, a subgroup of 34 participants recorded their dietary intake for 3 days using myfood24 (29). According to the recommended daily intakes (RDI) set by the Diabetes Care Programmes (45), carbohydrate and protein intake are both low and the fat intake is above recommendations. Of the 8 CGM metrics assessed, mean glucose and AUC showed significant associations with dietary mediators. Our exploratory analysis shows an increase in AUC and glucose levels associated with carbohydrate and fat intake. Various dietary carbohydrates – e.g. glucose, sucrose, cooked starches found in pastas and white bread) are readily digested and absorbed in the small intestines, this contributes to a rapid increase in blood glucose (46). Other studies have established that maternal glucose responses can be considerably influenced by the total amount of carbohydrates consumed (46). Increased dietary fat intake (high in saturated fat) has been associated with increased PPG levels and circulating fatty acids (47). Chronic increased level of circulating fatty acids have been linked to increased insulin resistance and inflammation, which are associated with risk of preeclampsia and preterm delivery (47, 48). Additionally, previous studies have demonstrated that elevated PPGRs contribute to an increased glucose transport to the foetus correlating with infant size and/or adiposity (46). Furthermore, our results showed that increasing protein intake by 1 standard deviation (while holding dietary carbohydrates and fats quantities constant) is associated with lower mean glucose and total AUC. While current positions and recommendations of major health bodies [National Health Services (UK), Canadian Diabetes Association, the American Diabetes Association, and the European Association for the Study of Diabetes] focus on replacing low-quality processed (high glycemic-index) carbohydrates with high-quality (low glycemic index) carbohydrates for diabetic patients, our analysis positions protein as an additional dietary pathway to manage gestational dysglycemia. The influence of protein on glycemia is likely to be explained by its more efficacious effect stimulating a rise in glucagon levels than glucose is in suppressing it – i.e. based on weight, protein is 10 times more efficacious than glucose in affecting the glucagon response in normal individuals (18). A previous study has concluded that substituting some of the fruit content with slowly digestible starch sources (e.g. legumes and al dente pasta, etc.), and increasing the protein content may result in a diet that is more acceptable for management of T2DM (49). Although this study was not designed to investigate interactions between carbohydrates quality consumed and time of day, future studies may be appropriately designed to investigate such an interaction and report on the importance of timing high nutritional-quality meals to manage dysglycemia.

This study has offered insight into temporal changes of dysglycemia and demonstrated the value of commonly reported CGM metrics, however, there are limitations to the study. First, although the study population was ethnically diverse (≈40% non-European ancestry), all women were diagnosed with GDM according to U.K. NICE criteria (3); therefore, our study population may not be representative of women diagnosed for GDM by alternative criteria (e.g., IADPSG – International Association of Diabetes and Pregnancy Study Group) (50, 51). Second, the CGM data were obtained at one time-period of gestation, which may not be representative of glycemia at other times during the pregnancy. Third, due to unequal number of total measurements between days and participants, we chose to average the 7-days data (that was available for participants) into a 24-hr period to analyse. While this prevented us from assessing a glucose shifts over multiple days or comparing weekdays and weekends, it allowed us to identify timepoints in a 24-hour period where glucose excursions were common. Finally, dietary logs were available only for a subgroup of participants and their mealtimes were not recorded; nonetheless, our analyses suggest future investigations of the role of dietary protein and carbohydrate quality on dysglycemia are warranted.

In summary, these results confirm that CGM is a rich source of information that could detect and quantify periods of dysglycemia. Additionally, we demonstrate that each of the metrics available to characterise CGM data, offers unique information to characterise an individual glucose profile and its variability. Therefore, demonstrating the complexity of maternal dysglycemia, which is not easily summarised by a single glycemic metric. Moreover, individuals assigned to diet with metformin appeared to have the greatest difficulty managing glycemia, suggesting the need for more directed care and follow-up may benefit this group of individuals. Finally, our exploratory analysis suggests that increased protein intake may assist with dysglycemia management, and that consideration of both protein and carbohydrate quality may provide optimal support for managing dysglycemia.

## Data Availability

All data produced in the present work are contained in the manuscript.

## Acknowledgements

The authors thank all the participants of the original study. The authors would also acknowledge the invaluable support from the diabetes antenatal care teams involved during original data collection.

## Author contributions

EMS designed the original study protocol. CFD, EMS and MAZ contributed to design of secondary analysis plan. EMS provided the CGM in GDM dataset. JDM provided the dietary data in the dataset. CFD and MAZ prepared the data for analysis. CFD, MAZ, JED, EMS, and MJH contributed to the data analysis and statistical analysis. CFD and MAZ have primary responsibility for the final content. CFD wrote the first draft of the manuscript. EMS, MDC, JED, and MJH provided critical feedback. CFD, MAZ, EMS, MDC, JED, and MJH read and approved the final manuscript. CFD and MAZ is the guarantor of this work and, as such, takes responsibility for the integrity of the data and the accuracy of the data analysis.

## Data sharing plan

Data described in the manuscript and analytic code will be made available upon request pending application and approval.

## Abbreviations

AUC: Area under the curve
BMI: Body Mass Index
CGM: Continuous glucose monitoring
CV: Coefficient of variation
GDM: Gestational diabetes mellitus
GI: Glycemic index
iAUC: Incremental area under the curve
NICE: National Institute for Health and Care Excellence
OR: Odds ratio
PPG: Postprandial glucose
PPGR: Postprandial glucose response
RDI: Recommended daily intakes
SD: Standard deviation
SMBG: Self-monitored blood glucose
T2DM: Type 2 diabetes mellitus
TAR: Time above range
TBR: Time below range
TIR: Time in range

