## Supplementary figures and images for "Observational assessments of the relationship of dietary and pharmacological treatment on continuous measures of dysglycemia over 24 hours in women with gestational diabetes"

### Supplementary Figure 1

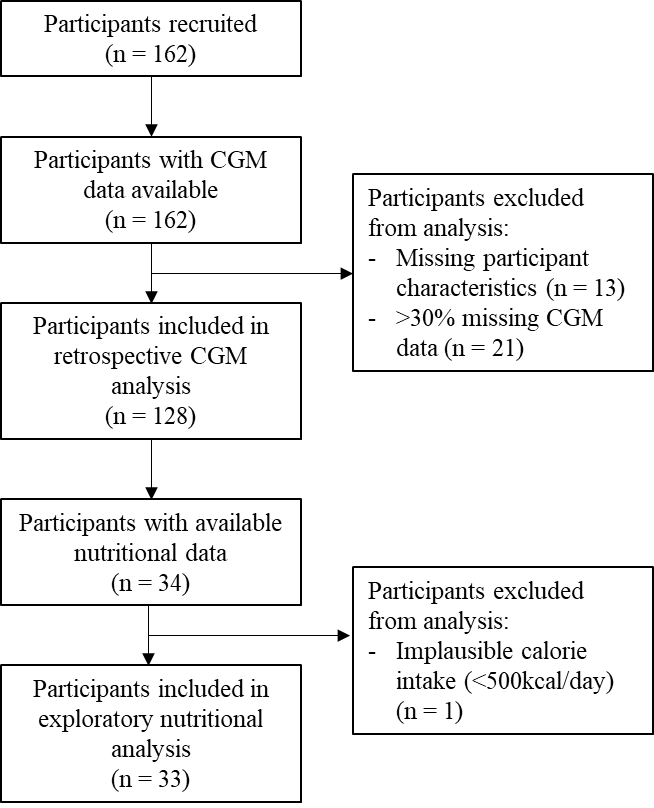

**Supplementary figure 1.** Participant flowchart
